# Awareness about the risk of hearing loss after ototoxic treatments in Swiss childhood cancer survivors

**DOI:** 10.1101/2024.09.23.24314181

**Authors:** Philippa Jörger, Carina Nigg, Maša Žarković, Grit Sommer, Martin Kompis, Gisela Michel, Marc Ansari, Nicolas Waespe, Claudia E. Kuehni

**Author notes:** **Corresponding author**: Correspondence to: Claudia E. Kuehni, Prof. MD; Institute of Social and Preventive Medicine, University of Bern, Mittelstrasse 43, 3012 Bern, Switzerland.

## Abstract

**Objectives:** The International Guideline Harmonization Group recommends childhood cancer survivors (CCS) exposed to ototoxic treatments be aware of the risk of hearing loss. We assessed awareness among adult CCS.

**Methods:** We identified adults diagnosed with cancer <20 years who received ototoxic treatments through the Swiss Childhood Cancer Registry (ChCR) and invited them to the HEAR-study. Participants completed a questionnaire and underwent pure-tone audiometry. Cancer and treatment data came from the ChCR. We used logistic regression to explore factors influencing awareness.

**Results:** 105 of 424 invited CCS participated (25%). 57% did not remember receiving information on hearing loss prior to the study. CCS who remembered being informed were more likely diagnosed after 1995 (OR: 4.5, 95% CI: 1.3-15.4), reported hearing problems (10.9, 2.6-45.1) and other late effects (4.1, 1.3-13.2), and treated with platinum chemotherapy only (10.8, 2.2-53.2) versus cranial radiotherapy only. 44% of participants presented clinically relevant hearing loss.

**Conclusions:** Over half of CCS exposed to ototoxic treatments were unaware of their risk of hearing loss.

**Practice Implications:** Educating CCS about potential late effects of ototoxic treatments is important to allow early diagnosis and treatment, especially for those who had cancer longer ago and those exposed to cranial radiation.

**Highlights:**

- Half of CCS exposed to ototoxic treatment were unaware of the risk of hearing loss
- CCS treated before 1995 were less aware about the risk of hearing loss
- Information provision is key to effective monitoring and early intervention

## 1. Introduction

Childhood cancer survivors (CCS) are at risk of hearing loss as a late effect of certain treatments [1]. Forty percent of CCS treated with platinum chemotherapy, around 20% of those treated with doses of cranial radiation ≥ 30 gray (Gy), and varying proportions of those treated with surgery involving the auditory system experience hearing loss [1–6]. Cumulative incidence of hearing loss due to these treatments increases up to 15 years after completing cancer treatment [7].

Among CCS, undetected hearing loss can impair language development, school performance, neurocognitive functions, and overall quality of life [8–11]. Regular audiometric screening enables hearing loss to be detected as early as possible and allows appropriate measures to be initiated. The International Guideline Harmonization Group (IGHG) recommends that CCS treated with a known ototoxic treatment (cisplatin with or without high-dose carboplatin (> 1500 mg/m²), and/or head or brain radiotherapy ≥ 30 Gy) and their health-care providers “should be aware of the risk of hearing loss” [1]. IGHG recommends annual hearing tests until six years of age, biannual tests until twelve years, and every five years thereafter [1].

Informing CCS about the risk for late effects related to their cancer treatment is important to facilitate follow-up care and to manage long-term health [12,13]. Previous research in Switzerland suggests CCS have limited knowledge about their risks for late effects [14–16]. Studies investigating information provision on specific late effects, such as hearing loss, are lacking.

Using data from the HEAR-study, a health service research project on the hearing of CCS, we first examined whether adult CCS who had received ototoxic treatments were aware of their risk for hearing loss, and had had their hearing evaluated as recommended. Second, we compared CCS who were aware of hearing loss risk with those who were not to identify CCS at increased need for information and support. Third, we investigated whether CCS currently experience clinically relevant hearing loss and explored the associations between awareness, hearing evaluation after end of treatment, and current hearing loss based on audiometric testing.

## 2. Methods

### 2.1 Study population

Eligible CCS were identified through the Swiss Childhood Cancer Registry (ChCR). The ChCR is a national registry that includes all Swiss residents diagnosed with leukemias, lymphomas, central nervous system (CNS) tumors, malignant solid tumors, or Langerhans cell histiocytosis before 20 years of age since 1976 [17,18]. We included CCS registered in the ChCR who had survived at least two years since diagnosis, had received ototoxic treatment as defined per IGHG guidelines, and were ≥ 18 years at the time of study. We excluded CCS who lacked a valid address, were living outside Switzerland, did not speak German or French, and had been contacted for another study in the past six months (January 2022-July 2022). The Ethics Committee of the Canton of Bern approved the ChCR (166/2014) and the HEAR-study (2021-01624).

### 2.2 Procedures and study design

The HEAR-study is a health service research project set up in 2021 at the University of Bern, Switzerland. Details have been published previously [19]. Eligible CCS were invited by postal mail; we sent up to three reminders to CCS who did not respond. CCS who responded with informed consent received an e-mail request to complete an online questionnaire (Supplementary Table S6) and an invitation to attend a free hearing test. Participants performed hearing tests at a local hearing aid shop where a certified acoustician performed bilateral pure-tone audiometry (125-8000Hz) with an additional bone conduction measurement (250-4000Hz) if the hearing threshold was > 25dB. We collected questionnaires and audiograms between July 15, 2022, and July 15, 2023.

### 2.3 Hearing-related information

#### 2.3.1 Awareness about the risk of hearing loss and information on prior hearing evaluations

We assessed awareness about the risk of hearing loss using a questionnaire. We asked participants if they had been informed about an increased risk of hearing loss (yes/no/don’t know). We also asked if they had been counseled to monitor their hearing after end of treatment (yes/no/don’t know). If participants answered yes to either of the two questions, we coded being informed as “yes”. Further, we asked if their hearing had ever been evaluated after end of treatment (yes/no/don’t know).

#### 2.3.2 Self-reported hearing loss, perceived hearing difficulties, and other late effects

We asked participants if they experienced any late effects of cancer or its therapy (yes/no), and to list them. We created a variable “late effects other than hearing loss” (yes/no) for which "no" indicated participants who only listed hearing loss or nothing. If participants listed hearing loss as a late effect in the open question, the variable on self-reported hearing loss was coded as “yes”. Further, we asked if they had problems following a conversation when there was background noise (hearing difficulties in noisy environments) (yes/no) and if they felt they did not hear well (feeling of poor hearing) (yes, often/yes, sometimes/no).

#### 2.3.3 Hearing loss from audiogram measurements

The hearing aid shops sent us audiogram results in a coded file. For grading we visualized the audiograms using the R package “audiometry” (Lehnbert B, R package version 0.3.0, 2021) in RStudio (R Core Team, Vienna, AUT, 2022). Two trained researchers (PJ and CN) independently graded audiograms for each ear according to the SIOP Boston Ototoxicity Scale [20], an established grading scheme used for CCS exposed to ototoxic treatments [21,22]. We resolved discrepancies through discussions with an experienced audiologist (MK). We considered bone conduction measurements when there was consistently a significant difference between air and bone conduction measurements (≥ 10 dB), indicating a potential conductive hearing loss [20]. We used the grading of the more affected ear. Severity was graded as (a) none (grade 0), (b) mild (grade 1), (c) moderate (grade 2), or (d) severe hearing loss (≥ grade 3) [20]. Clinically relevant hearing loss was defined as SIOP Boston grade ≥ 2 [20,22].

### 2.4 Participant characteristics

#### 2.4.1 Clinical and cancer-treatment related characteristics

The ChCR provided data on biological sex, year of birth, cancer diagnosis, age at diagnosis, year of diagnosis, and type of therapy including carboplatin chemotherapy, cisplatin chemotherapy, and cranial radiation.

#### 2.4.2 Information on follow-up care

We asked participants in the questionnaire if they were still undergoing clinical follow-up care related to their cancer (yes/no), and if they had received any information regarding follow-up care in general (yes/no/don’t know).

#### 2.4.3 Socioeconomic characteristics

The questionnaire assessed sociodemographic characteristics including migration background (yes/no), participants’ and their parents’ education levels, and language region (German/French). Participants were categorized as having a migration background if at least one parent was born outside Switzerland. We defined three education level categories: primary schooling (compulsory schooling only, ≤ 9 years), secondary education (vocational training, upper secondary education), and tertiary education (university, or technical college).

### 2.5 Statistical analysis

First, we assessed how many CCS were aware of a risk of hearing problems (information status) and how many had completed a hearing evaluation after the end of treatment. For further analysis, we combined the categories “no” and “don’t know” interpreting both as insufficient awareness. Next, we assessed severity of hearing loss and prevalence of clinically relevant hearing loss (SIOP Boston Grade ≥ 2). Using univariable logistic regression, we investigated factors associated with information status and having completed a hearing evaluation after end of treatment (Supplementary Tables S4 and S5). We included sex, age at study, time since diagnosis, age at diagnosis, year of diagnosis, language region, participant and parental education, migration background, presence of late effects other than hearing loss, self-reported hearing loss, general information received on follow-up care, attending regular follow-up care visits, and treatment (only platinum/only cranial radiation ≥ 30 Gy/both platinum and cranial radiation) as explanatory variables. We categorized year of diagnosis into the periods before and after 1995, reflecting a cut-off after which a decline in hearing loss was observed in previous studies of CCS from Switzerland [7]. Information status was included as an additional explanatory variable for the outcome hearing evaluation. In a multivariable logistic regression, we included all exposures associated with the outcome at a significance level of p ≤ 0.1. If there were missing values in independent or dependent variables, we performed complete case analyses.

## 3. Results

### 3.1 Study population

Of 573 eligible CCS exposed to ototoxic treatments, identified through the ChCR, 424 were invited to the study. One-hundred and five (25%) consented and completed the questionnaire and 83 (20%) also completed a hearing test as part of our study (Supplementary Figure SF1). About half (n=57, 54%) of participants were female, median age at study start was 32 years (interquartile range [IQR]: 24-37), age at diagnosis 8 years (IQR: 4-13), and median time since diagnosis 23 years (IQR: 19-32) (Table 1). Common diagnoses were malignant bone tumors (n=24, 23%), CNS tumors (n=23, 22%) and soft tissue sarcomas (n=22, 21%). Eighty CCS (76%) had been treated with platinum chemotherapy, and 41 (39%) with cranial radiation (Supplementary Table S2). Compared to nonparticipants, CCS completing the questionnaire were more often female, older at time of study, and further away from diagnosis (Supplementary Table S1).

**TABLE 1.**
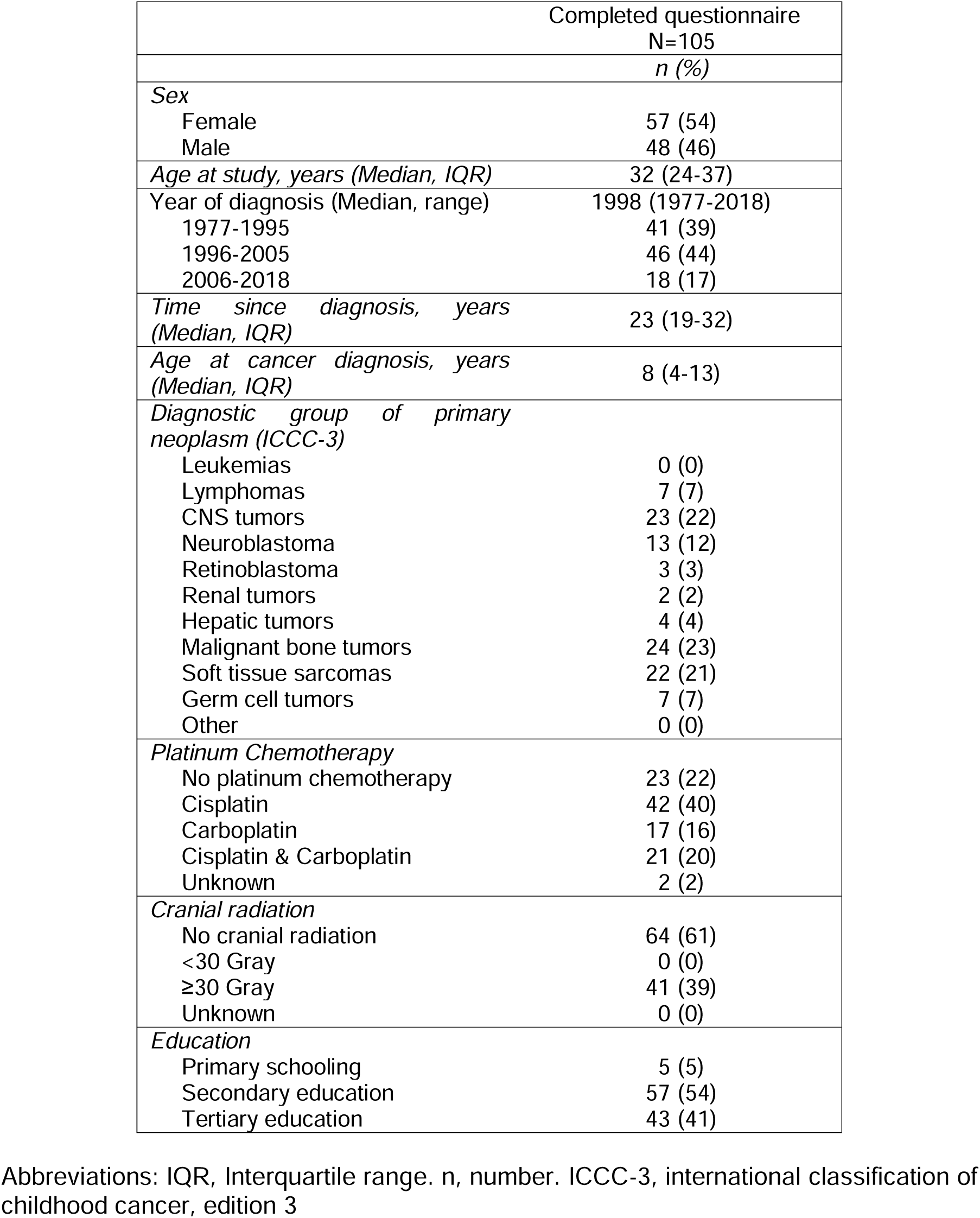
Characteristics of participants completing the questionnaire (N=105).

### 3.2 Information status and prior hearing evaluation

Among 105 participants, 46 (44%, 95% confidence interval [CI]: 35-54%) reported having received information or advice regarding risk of hearing loss or necessity of conducting hearing tests after end of treatment, while 31 (30%, 95% CI: 22-39%) reported not having received information/advice and 28 (27%, 95% CI: 19-36%) did not remember (Figure 1). Similarly, 40 (38%, 95% CI: 29-48%) participants reported having completed a hearing evaluation after end of treatment, 34 (32%, 95% CI: 24-42%) reported not having completed a hearing evaluation, and 31 (30%, 95% CI: 22-39%) did not remember. Of the 40 participants reporting a hearing evaluation, 13 (33%) had done it in the last 5 years, 12 (30%) longer ago, and 15 (38%) did not remember the year of their last hearing evaluation (Supplementary Table S3).

**FIGURE 1.**
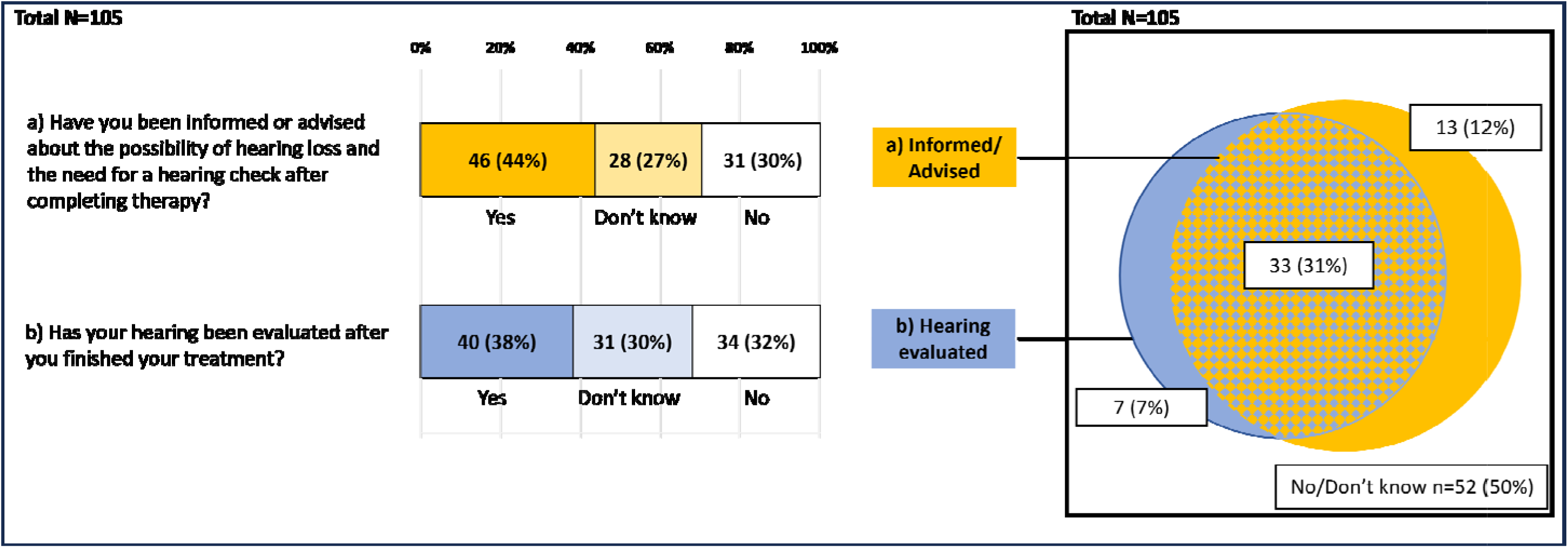
Participant responses to questions on awareness and prior hearing evaluation. Participants who reported (a) having been informed or advised about the possibility of hearing loss and the need for hearing tests after completing erapy and (b) having had a hearing evaluation after completing the therapy. Proportional Venn-Diagram depicting the relation between having en informed or advised and having had a hearing evaluation. Numbers represent the number (n) and percentage (%) of participants reporting yes to the respective question; (a) Informed/Advised, Were you formed about hearing loss as a potential late effect of your treatment / Have you been advised to have your hearing checked after completing your therapies? (b) Hearing evaluated, Has your hearing ever been checked after you finished your treatment? No/Don’t know; Answered no or do t know to both questions. Abbreviations: number.

We visualized the relationship between information status and prior hearing evaluation using a proportional Venn diagram (Figure 1). Most CCS who were informed also had their hearing evaluated at some point after treatment.

### 3.3 Degree of hearing loss and information status, self-reported hearing loss, and prior hearing evaluation

Of 83 participants who completed a hearing test for the study, 29 had normal hearing (35%, 95% CI: 25-46), 17 (20%, 95% CI: 13-31%) had mild, 11 (13%, 95% CI: 7-23%) moderate, and 26 (31%, 95% CI: 22-42%) severe hearing loss, resulting in 37 participants (45%, 95% CI: 34-56%) with clinically relevant hearing loss. Among the 36 participants with clinically relevant hearing loss who completed the questionnaire, 20 (56%, 95% CI: 39-71%) reported having received information/advice regarding their risk for hearing loss and 14 (39%, 95% CI: 24-56%) reported having had their hearing evaluated after end of treatment. Further, 19 (53%, 95% CI: 36-69%) of the participants with clinically relevant hearing loss reported prior hearing problems in the questionnaire, 29 (81%, 95% CI: 64-91%) reported hearing difficulties in noisy environments, and 30 (83%, 95% CI: 67-93%) sometimes or often felt like they were not hearing well. (Figure 2)

**FIGURE 2.**
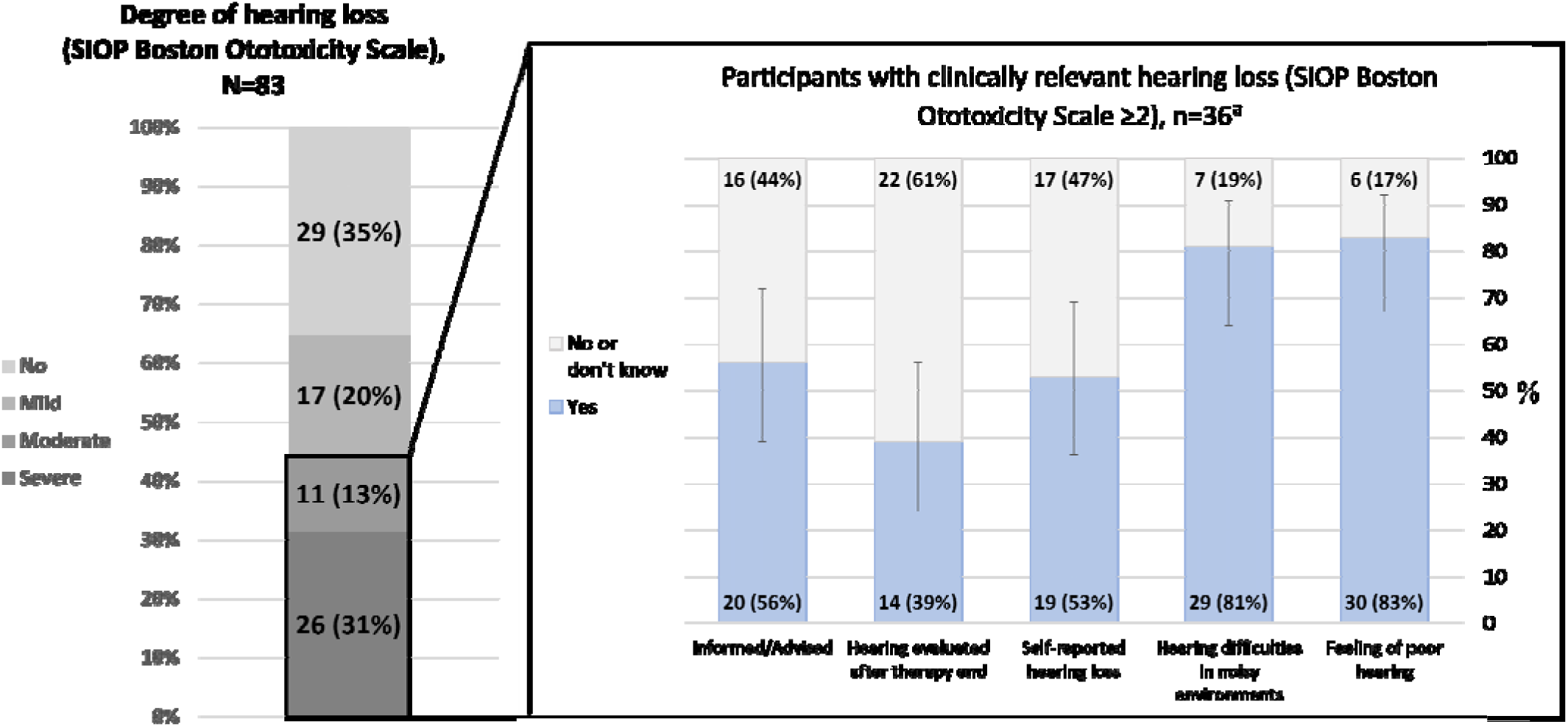
Degree of hearing loss assessed with audiograms and graded according to SIOP Boston ototoxicity scale, and responses of questions on awareness with clinically relevant hearing loss (Grade≥2). (left) Degree of hearing loss according to SIOP Boston Ototoxicity Scale for participants with available audiograms, N=83. No hearing loss, grade 0. Mild, grade 1. Moderate, grade 2. Severe, ≥ grade 3. (right) Participants with clinically relevant hearing loss (SIOP Boston ototoxicity scale ≥2, n=36) who completed the baseline questionnaire and status on information/advice received on hearing loss (yes/no), hearing evaluated after therapy end (yes/no), self-reported hearing loss (yes/no), hearing difficulties in noisy environments (yes/no) and feeling of bad hearing (yes/no). 95% Confidence intervals are displayed for those reporting “Yes”. ^a^, Participants with clinically relevant hearing loss who did not complete the baseline questionnaire were excluded from this graphic (n=1). Abbreviations: SIOP, International Society of Paediatric Oncology. n, number.

### 3.4 Factors associated with information status and hearing evaluation after end of treatment

Participants informed about risk of hearing loss were diagnosed more often after 1995 (odds ratio [OR]: 4.5, 95% CI: 1.3-15.4), reported more prior hearing problems (OR: 10.9, 95% CI: 2.6-45.1) and other late effects (OR: 4.1, 95% CI: 1.3-13.2), and were more often treated with platinum chemotherapy (OR: 10.8, 95% CI: 2.2-53.2) or both platinum chemotherapy and cranial radiotherapy (OR: 6.5, 95% CI: 1.0-41.8) compared to cranial radiotherapy alone (Table 2). Those who were informed were more likely to have had their hearing evaluated after end of treatment (OR: 13.6, 95% CI: 3.8-48.3).

**TABLE 2.**
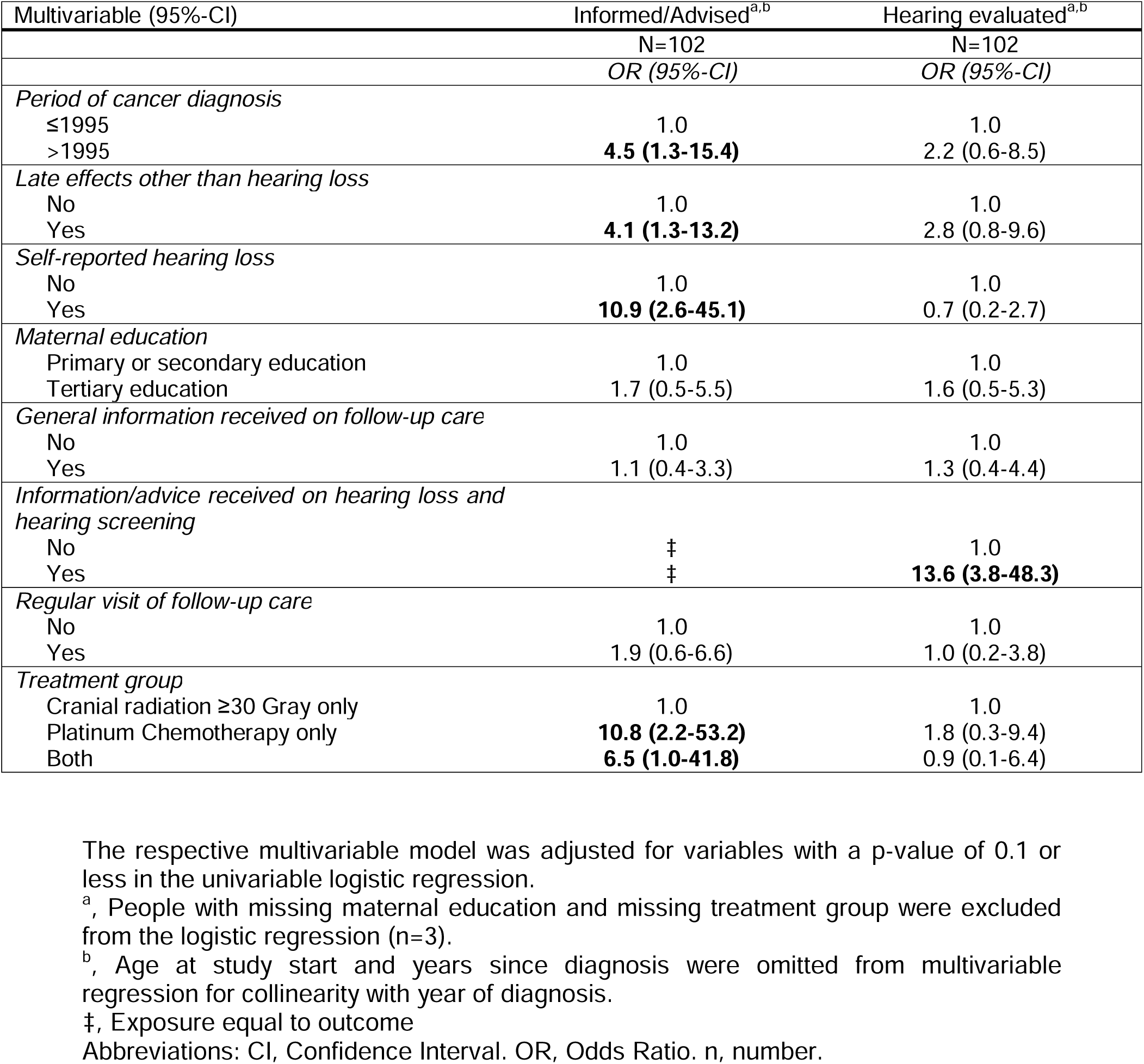
Factors associated with a) having received information/advice about hearing loss, and b) having ever completed a hearing evaluation after therapy; results from multivariable regressions.

| Multivariable (95%-CI) | Informed/Advised<sup>a,b</sup> | Hearing evaluated<sup>a,b</sup> |
| --- | --- | --- |
|  | N=102 | N=102 |
|  | OR (95%-CI) | OR (95%-CI) |
| *Period of cancer diagnosis* |  |  |
| ≤1995 | 1.0 | 1.0 |
| >1995 | **4.5 (1.3-15.4)** | 2.2 (0.6-8.5) |
| *Late effects other than hearing loss* |  |  |
| No | 1.0 | 1.0 |
| Yes | **4.1 (1.3-13.2)** | 2.8 (0.8-9.6) |
| *Self-reported hearing loss* |  |  |
| No | 1.0 | 1.0 |
| Yes | **10.9 (2.6-45.1)** | 0.7 (0.2-2.7) |
| *Maternal education* |  |  |
| Primary or secondary education | 1.0 | 1.0 |
| Tertiary education | 1.7 (0.5-5.5) | 1.6 (0.5-5.3) |
| *General information received on follow-up care* |  |  |
| No | 1.0 | 1.0 |
| Yes | 1.1 (0.4-3.3) | 1.3 (0.4-4.4) |
| *Information/advice received on hearing loss and hearing screening* |  |  |
| No | ‡ | 1.0 |
| Yes | ‡ | **13.6 (3.8-48.3)** |
| *Regular visit of follow-up care* |  |  |
| No | 1.0 | 1.0 |
| Yes | 1.9 (0.6-6.6) | 1.0 (0.2-3.8) |
| *Treatment group* |  |  |
| Cranial radiation ≥30 Gray only | 1.0 | 1.0 |
| Platinum Chemotherapy only | **10.8 (2.2-53.2)** | 1.8 (0.3-9.4) |
| Both | **6.5 (1.0-41.8)** | 0.9 (0.1-6.4) |

The respective multivariable model was adjusted for variables with a p-value of 0.1 or less in the univariable logistic regression.
<sup>a</sup>, People with missing maternal education and missing treatment group were excluded from the logistic regression (n=3).
<sup>b</sup>, Age at study start and years since diagnosis were omitted from multivariable regression for collinearity with year of diagnosis.
‡, Exposure equal to outcome
Abbreviations: CI, Confidence Interval. OR, Odds Ratio. n, number.

## 4. Discussion and Conclusion

### 4.1 Summary of findings

This study found that over half of CCS who received ototoxic treatment reported they were unaware of the risk of hearing loss even though international guidelines recommend that they be aware. More than half did not remember having ever had their hearing evaluated after end of treatment. We found a strong association between being informed and having had a hearing evaluation. Almost half of participants had clinically relevant hearing loss assessed by pure-tone audiometry, yet almost half of those did not report hearing problems in the questionnaire, which suggests they were not aware of their hearing loss.

### 4.2 Information status

Previous surveys on information provision and information needs among Swiss CCS show that approximately one-third of survivors do not remember having ever received general information about potential late effects of their cancer or therapy, and about 80% of CCS wish to be better informed [14–16]. In this study, 56% of CCS did not remember receiving information about the risk for hearing loss. This is comparable to a Norwegian study about information provision in which up to 60% of cancer survivors overall did not recall receiving information on late effects including hearing loss, though the study also included survivors without ototoxic treatments [12]. A study from Hong Kong assessing knowledge about risk-based, exposure-related late effects found that, similar to our study, almost 60% of CCS were not aware of potential hearing problems following ototoxic treatments [13].

#### 4.2.1 Associations with information status

Consistent with other studies [12,14,15], we found that CCS seem to be less well informed the more time has passed since diagnosis. A reason for this might be that parents of patients who were younger children were the primary recipients of information [23]. Parents might not have passed on the information to their children, or CCS may have forgotten it given a long median time of 22 years since diagnosis.

We found that CCS diagnosed after 1995 were better informed, which is likely due to advances in knowledge about long-term effects of ototoxic treatments. For example, the American Speech-Language-Hearing Association (ASHA) guideline from 1994 recommended monitoring the hearing at least until one year post-treatment with platinum agents and only longer if hearing loss was detected [24]. Considering that one-third of our study population was diagnosed before 1995, they likely received outdated or limited information about potential long-term effects.

The 1994 ASHA guideline further stated that cranial radiation alone has little effect on hearing [24], and many clinicians might be less aware of the effect of cranial radiation on hearing than platinum chemotherapy. This might explain why participants who received cranial radiation ≥ 30 Gy alone were less informed than participants treated with platinum agents. This aligns with a previous record-based study in which we found that CCS treated with cranial radiation ≥ 30 Gy were less likely to have received post-therapy hearing evaluations than CCS treated with platinum chemotherapy [25]. Participants reporting other late effects were more informed about the risk of hearing loss, suggesting that CCS with multiple late effects are generally more aware of treatment-related complications [26].

### 4.3 Hearing evaluations after end of treatment

A retrospective, record-based study examining audiological evaluations post-therapy in Switzerland found that 28% of CCS at risk for hearing loss had not had any hearing evaluations after end of treatment [25]. This is comparable to the 32% observed in our study. In our study, a further 30% did not recall undergoing any hearing evaluations. While they might have had an initial test, CCS may not have had subsequent assessments when their hearing test result was normal as specified in older ASHA and Children’s Oncology Group Long-Term Follow-Up Guidelines [24,27]. Given that initial hearing tests often occur during or soon after treatment, many may have forgotten about them or were not contacted for later hearing evaluations as currently recommended [1].

### 4.4 Degree of hearing loss and self-reported hearing loss

We previously reported that 36% of CCS had clinically relevant hearing loss after platinum-based therapy according to the SIOP Boston ototoxicity scale [28]. This new study finds a higher prevalence (47%) of clinically relevant hearing loss, likely because of different characteristics of the study population, which now is older and further from diagnosis and more recently represents the survivor cohort. Considering the newer study’s low response rate, selection bias could have also played a role in the difference.

Almost half of our study participants with clinically relevant hearing loss did not report hearing problems as late effects in the questionnaire, suggesting they were not aware of them. The proportion of participants with hearing problems by SIOP Boston grade who did not self-report hearing loss was higher than that found in a previous study in which we compared questionnaire-reported hearing loss with audiometry results [29]. In that study, only 14% of CCS who did not self-report hearing loss had hearing loss in the SIOP-Boston grade range from 1 to 4. In the current study, self-reported hearing loss was assessed using an open question about known late effects. We have potentially underestimated the proportion of participants who are aware about their hearing loss since some participants with hearing loss may not have listed it as a late effect.

### 4.5 Strengths and limitations

A strength of this study is the population-based sample with comprehensive cancer-related medical data provided by the Swiss ChCR. Combined with the questionnaire-based information and recent audiograms, our dataset allowed for a thorough analysis. Study limitations include the response rate of 25% which, as noted above, potentially introduced a selection bias. However, although analyses revealed some statistically significant differences between responders and nonresponders regarding cancer-related characteristics, differences were modest based upon effect sizes. The low response rate may be due to the study settings as CCS had not only to return a questionnaire but also visit a hearing aid shop for a hearing test.

### 4.6 Conclusion

A large proportion of CCS are not aware of the potential risk of hearing loss due to their treatment. Among those who are aware of this late effect, most have undergone hearing evaluations. From this we conclude that knowledge is key to timely interventions that can mitigate the effects of hearing loss. Caregivers, parents, and CCS should always have the latest information on late effects and follow-up care at hand to manage and protect auditory health throughout the lives of CCS—even those with decades intervening since treatment.

### 4.7 Practice implications

How do we best manage survivorship to follow up treatment and address potential late effects? In addition to direct, clinical support of CCS and parents, electronic patient records continue to evolve and will improve the retention and support the acquisition of knowledge accessible by both providers and patients. A specific proposal for enhancing information access and retention for CCS is the Survivorship Passport, or SurPass [30]. This tool can provide crucial details about a survivor’s medical history and a tailored plan for follow-up to caregivers, and information for CCS that can inform their own decision-making; accessible online, it can be continually updated by new research findings. Highly secure yet accessible electronic records such as the SurPass can sustain awareness among caregivers and CCS wherever their journey through survivorship takes them.

## Data Availability

All data produced in the present study are available upon reasonable request to the authors

## 5. Abbreviations

ASHA: American Speech-Language-Hearing Association
CCS: Childhood cancer survivor
COG: Children’s Oncology Group
CI: Confidence interval
IQR: Interquartile range
Gy: Gray

## Acknowledgements

We thank the study team of the Childhood Cancer Research Group, Institute of Social and Preventive Medicine, University of Bern, the Swiss Childhood Cancer Survivor Study, and the team of the Swiss Childhood Cancer Registry. We thank Christopher Ritter from the editorial service of the Institute of Social and Preventive Medicine at the University of Bern for the editorial suggestions. We thank all childhood cancer survivors participating in the study.

## 6. CRediT authorship contribution statement

**Philippa Jörger:** formal analysis, investigation, writing—original draft, review & editing, visualization, project administration. **Carina Nigg:** formal analysis, writing—original draft, review & editing, supervision. **Maša Žarkovi**ć: formal analysis, writing—original draft, review & editing. **Grit Sommer:** writing—review & editing. **Martin Kompis:** formal analysis, writing—review & editing, funding acquisition. **Gisela Michel:** writing—review & editing, funding acquisition. **Marc Ansari**: conceptualization, writing—review & editing, supervision, funding acquisition. **Nicolas Waespe:** writing—review & editing, funding acquisition. **Claudia Kuehni:** conceptualization, methodology, formal analysis, writing— review & editing, supervision, project administration, funding acquisition.

---

**Supplementary FIGURE SF1.**
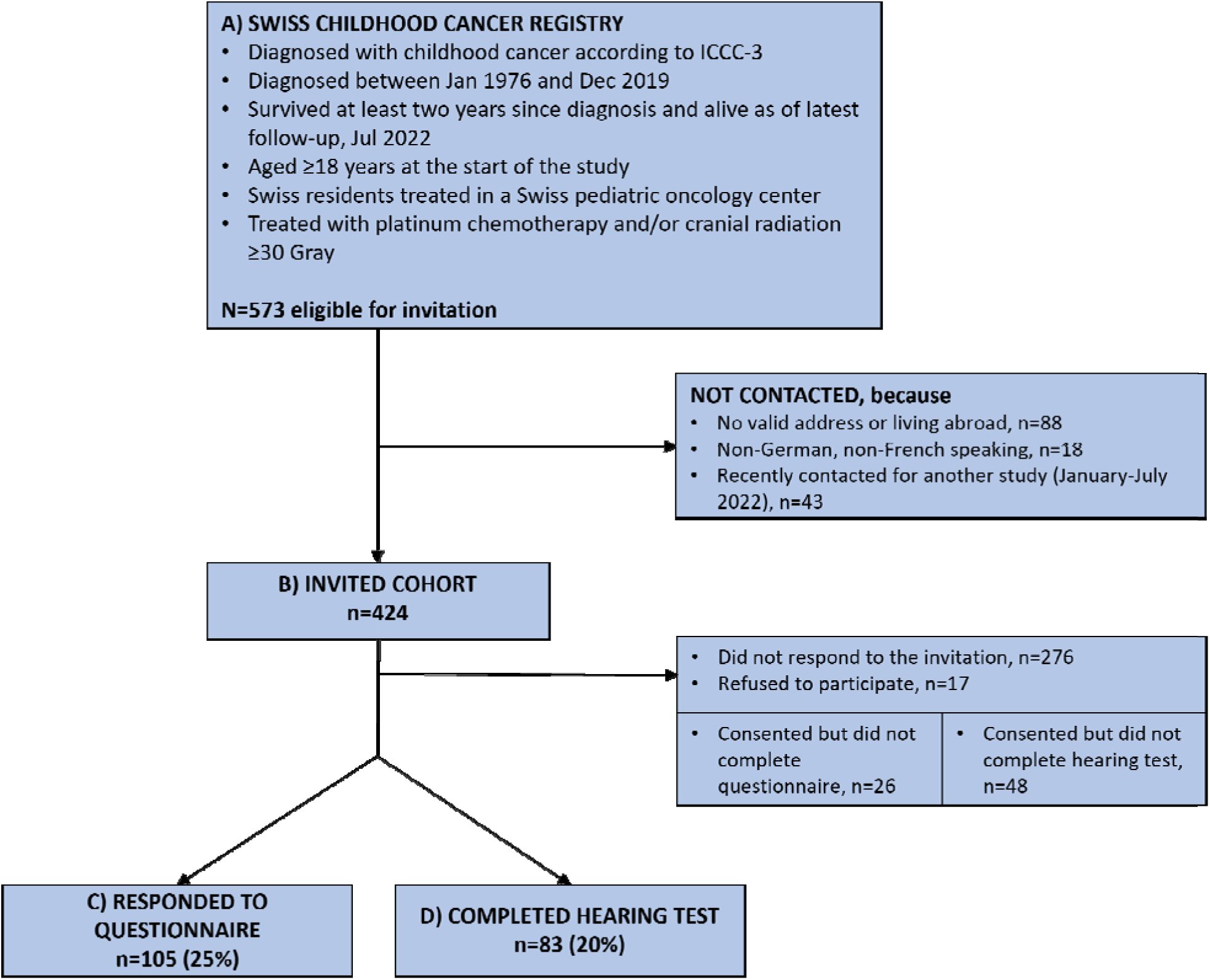
Flowchart illustrating the HEAR-study population, depicting eligible survivors identified by the Swiss Childhood Cancer Registry. Abbreviations: ICCC-3, international classification of childhood cancer, edition 3. n, number.

**Supplementary TABLE S1.**
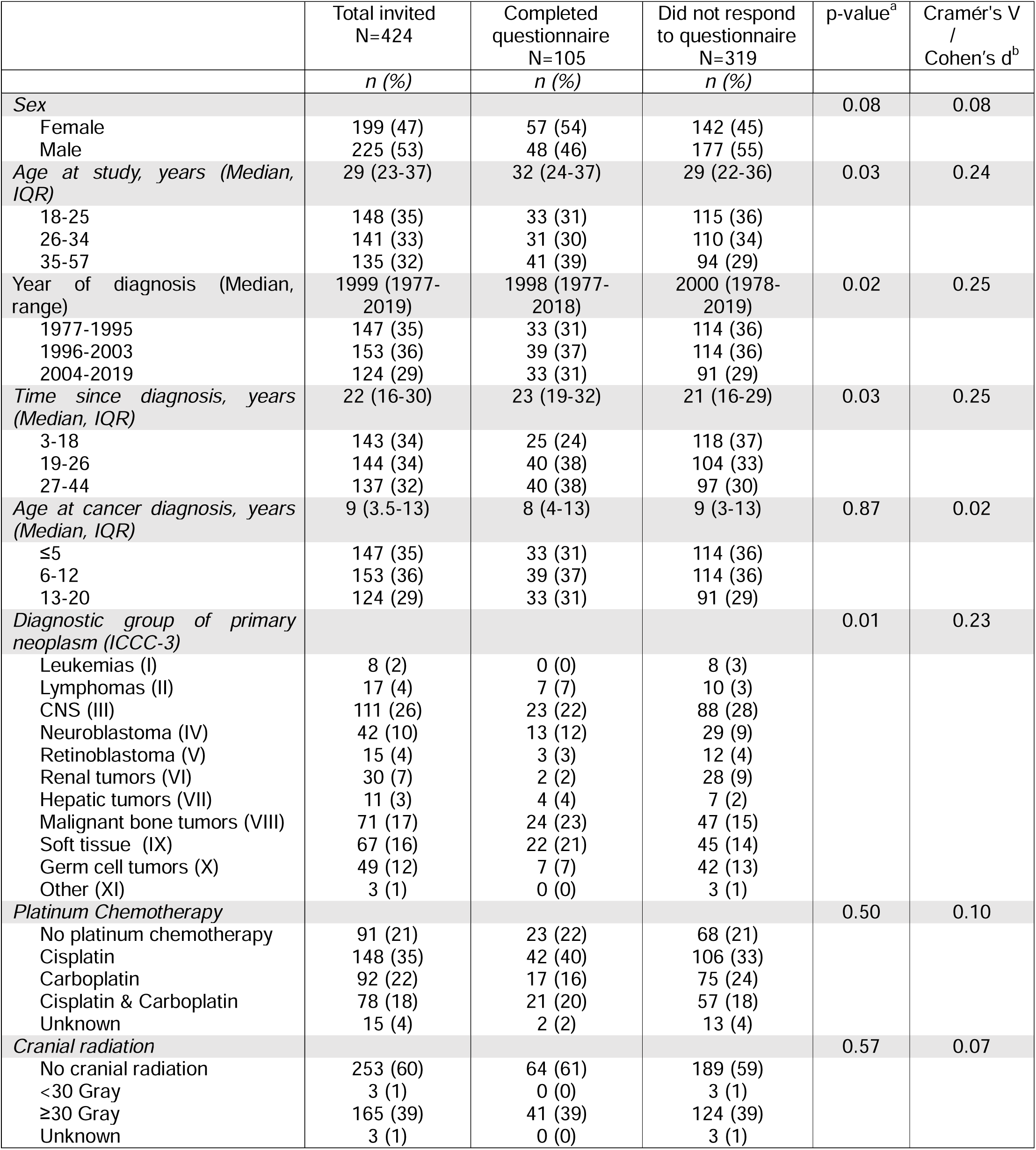
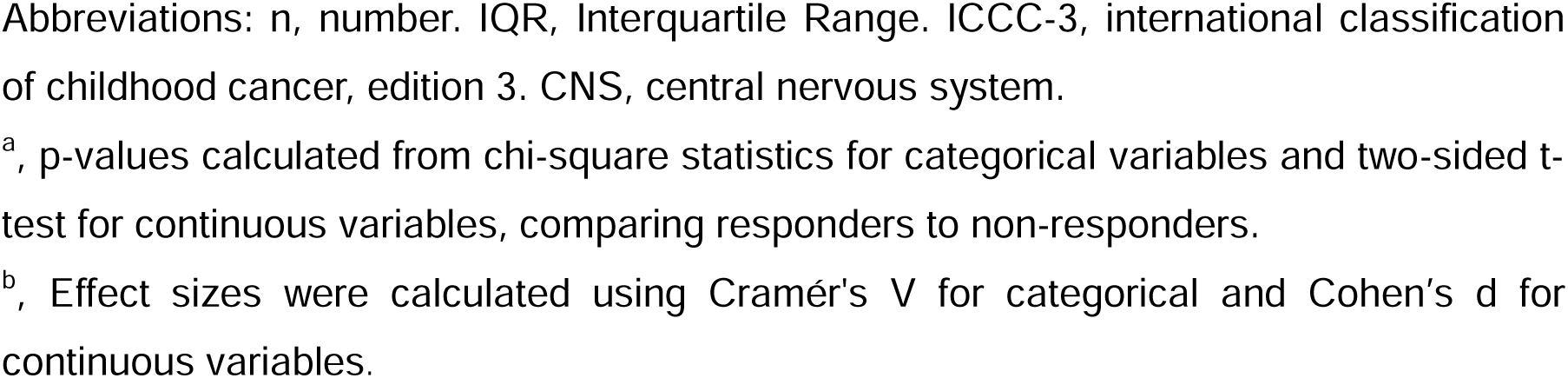
Demographic and clinical characteristics of study population, stratified by response status.

**Supplementary TABLE S2.** Clinical risk factors for hearing loss in childhood cancer survivors who participated in the HEAR-study (N=105).

|  | **Cranial radiation ≥30 Gray**<br>**n (%)** | **No cranial radiation**<br>**n (%)** | **Total**<br>**n (%)** |
| --- | --- | --- | --- |
| **Cisplatin** | 4 (10) | 38 (59) | 42 (40) |
| **Carboplatin** | 7 (17) | 10 (16) | 17 (16) |
| **Cisplatin & Carboplatin** | 6 (15) | 15 (23) | 21 (20) |
| **No platinum chemotherapy** | 23 (56) | 0 (0) | 23 (22) |
| **Unknown** | 1 (2) | 1 (2) | 2 (2) |
| **Total** | 41 (100) | 64 (100) | 105 (100) |

**Supplementary TABLE S3.** Calendar period of last completed hearing test (n=40).

| **Year** | **Participants N=40 (n, (%))** |
| --- | --- |
| <2020 | 12 (30) |
| 2020-2022 | 13 (33) |
| Do not remember | 15 (38) |

Abbreviations: n, number.

**Supplementary TABLE S4.**
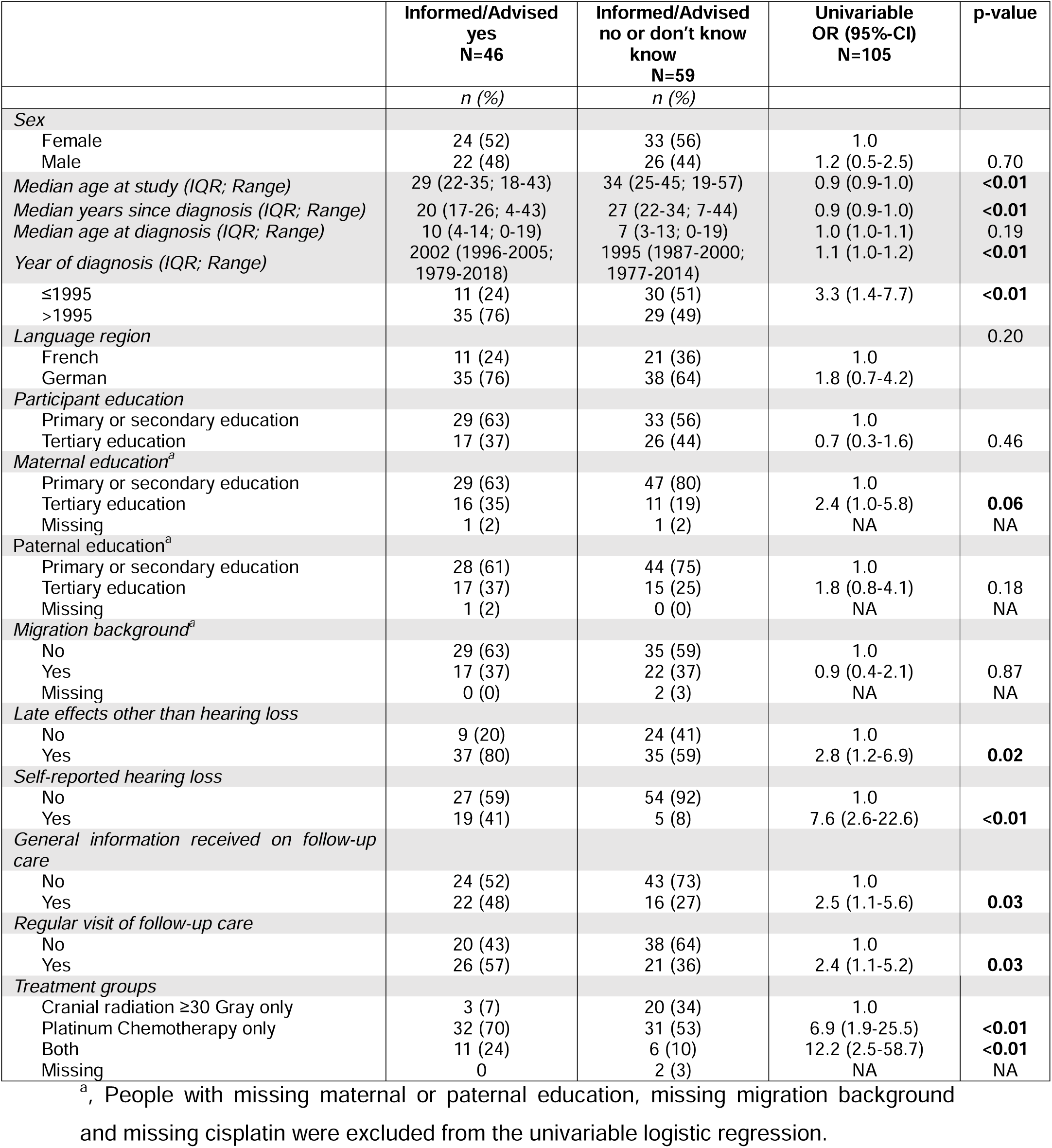
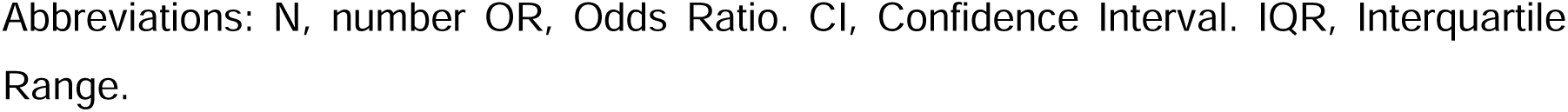
Characteristics of participants by information status and factors associated with information status; results from univariable logistic regressions.

**Supplementary TABLE S5.**
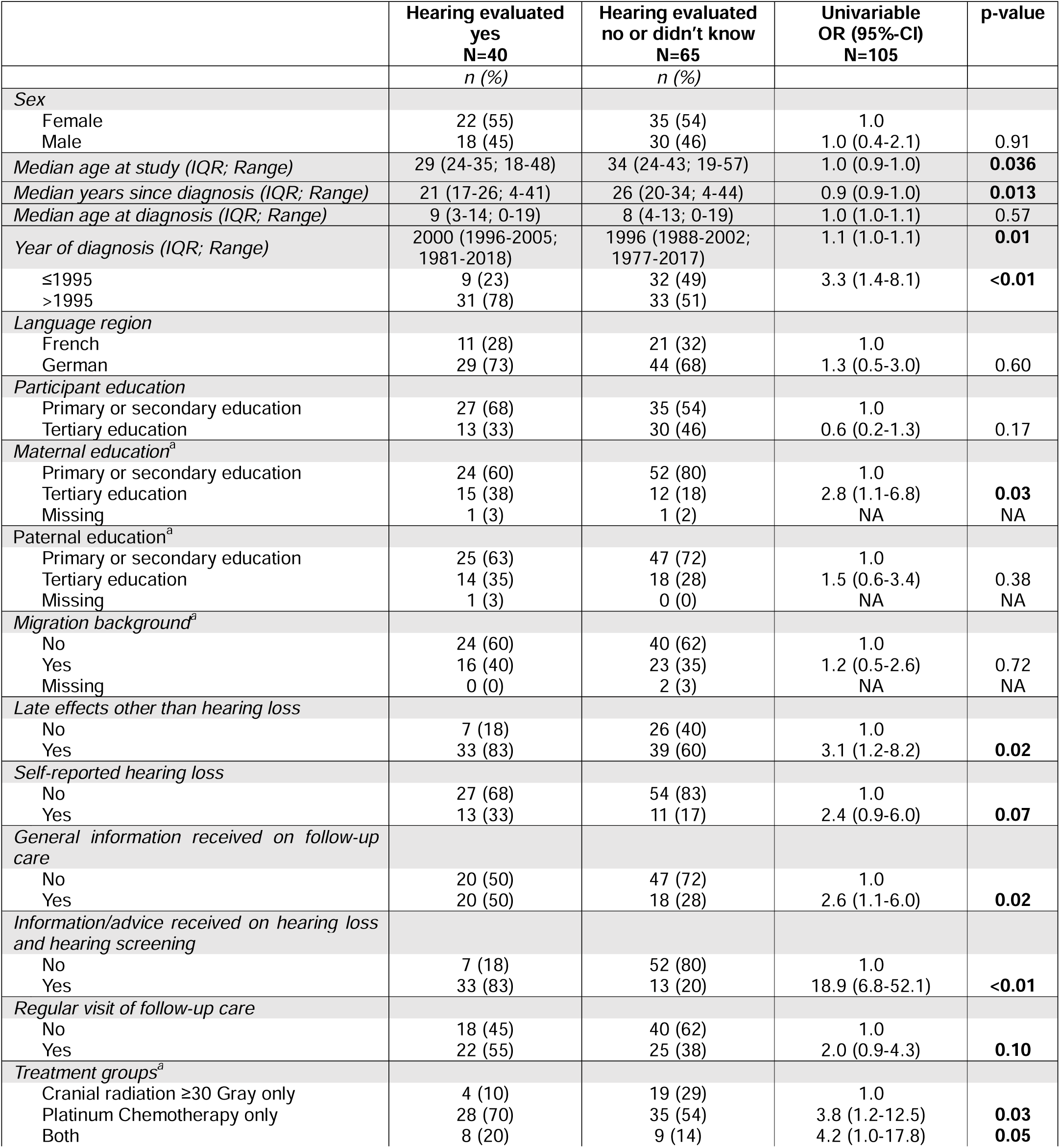
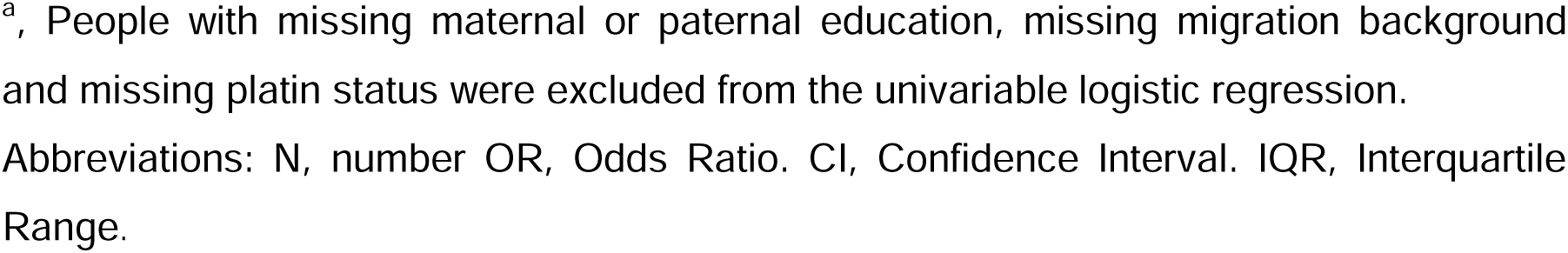
Characteristics of participants by status of prior hearing evaluation; results from univariable logistic regressions.

**Supplementary TABLE S6.**
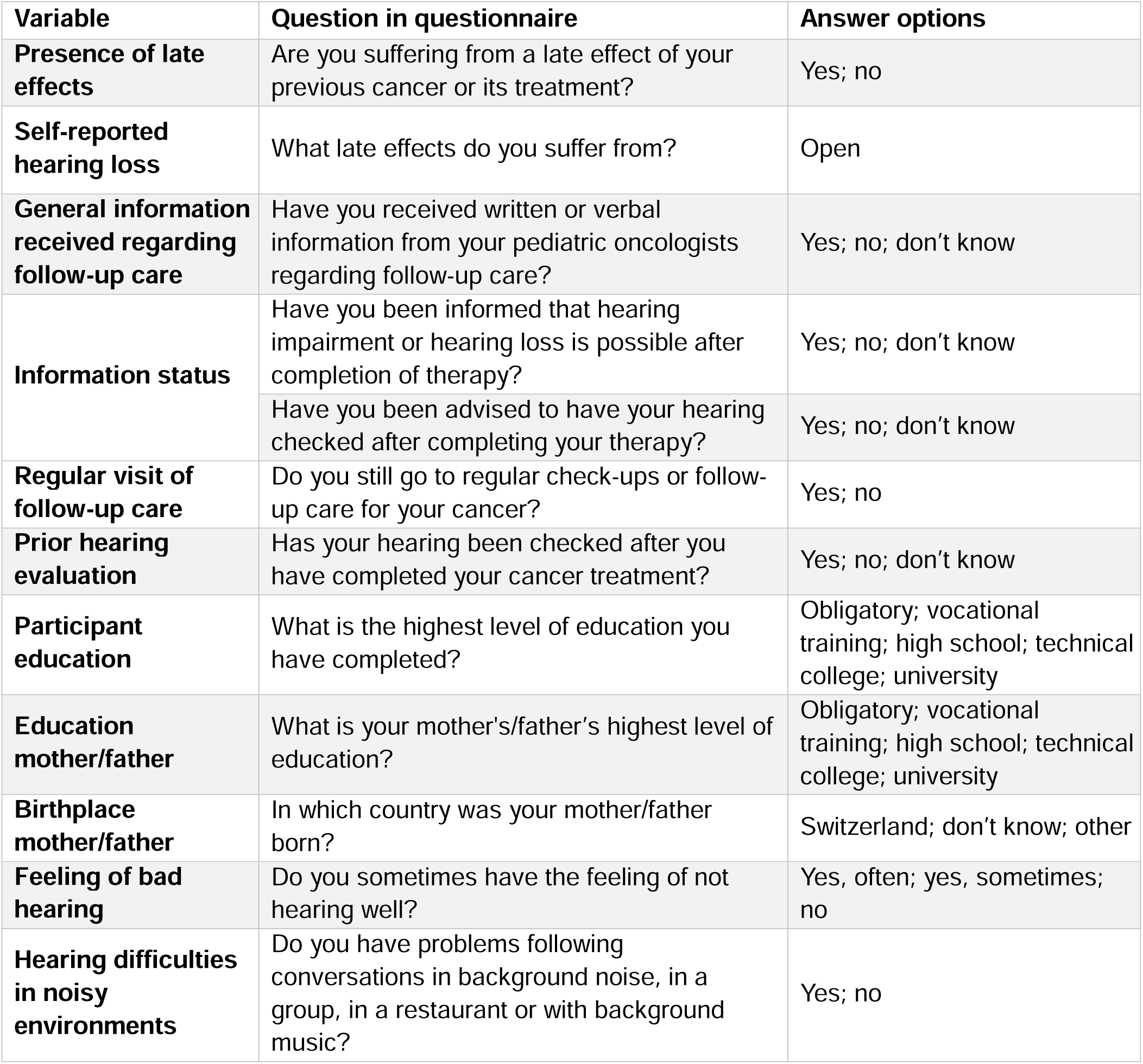
Questions on late effects, hearing loss, information provision, awareness of hearing loss, and sociodemographic characteristics in from the HEAR-study (translated into English).

| Variable | Question in questionnaire | Answer options |
| --- | --- | --- |
| **Presence of late effects** | Are you suffering from a late effect of your previous cancer or its treatment? | Yes; no |
| **Self-reported hearing loss** | What late effects do you suffer from? | Open |
| **General information received regarding follow-up care** | Have you received written or verbal information from your pediatric oncologists regarding follow-up care? | Yes; no; don't know |
| **Information status** | Have you been informed that hearing impairment or hearing loss is possible after completion of therapy? | Yes; no; don't know |
|  | Have you been advised to have your hearing checked after completing your therapy? | Yes; no; don't know |
| **Regular visit of follow-up care** | Do you still go to regular check-ups or follow-up care for your cancer? | Yes; no |
| **Prior hearing evaluation** | Has your hearing been checked after you have completed your cancer treatment? | Yes; no; don't know |
| **Participant education** | What is the highest level of education you have completed? | Obligatory; vocational training; high school; technical college; university |
| **Education mother/father** | What is your mother's/father's highest level of education? | Obligatory; vocational training; high school; technical college; university |
| **Birthplace mother/father** | In which country was your mother/father born? | Switzerland; don't know; other |
| **Feeling of bad hearing** | Do you sometimes have the feeling of not hearing well? | Yes, often; yes, sometimes; no |
| **Hearing difficulties in noisy environments** | Do you have problems following conversations in background noise, in a group, in a restaurant or with background music? | Yes; no |

